# Risk factors for labour induction and augmentation: a multicentre prospective cohort study in India

**DOI:** 10.1101/2023.12.21.23300361

**Authors:** Tuck Seng Cheng, Farzana Zahir, Carolin Solomi V, Ashok Verma, Sereesha Rao, Saswati Sanyal Choudhury, Gitanjali Deka, Pranabika Mahanta, Swapna Kakoty, Robin Medhi, Shakuntala Chhabra, Anjali Rani, Amrit Bora, Indrani Roy, Bina Minz, Omesh Kumar Bharti, Rupanjali Deka, Charles Opondo, David Churchill, Marian Knight, Jennifer J Kurinczuk, Manisha Nair

## Abstract

**OBJECTIVE:** To investigate clinical and non-clinical factors influencing labour induction and augmentation in pregnant women in India.

**DESIGN:** Prospective cohort study of 9305 pregnant women.

**SETTING:** 13 tertiary and community hospitals in six states across India.

**PARTICIPANTS:** Women ≥18 years of age and planning a vaginal birth in the study hospital were recruited in the third trimester of pregnancy (≥28 weeks of gestation) and followed-up during labour and up to 48 hours of childbirth.

**MAIN OUTCOME MEASURES:** Outcomes were induction and augmentation of labour as per childbirth records. Maternal and fetal clinical conditions in current pregnancy were abstracted from medical records at recruitment and after childbirth, and classified based on guidelines to generate induction-related clinical indication groups: (i) ≥2 indications, (ii) one indication, (iii) no indication and (iv) contraindication. Non-clinical factors included self-reported maternal socio-demographic and lifestyle factors, and maternal medical and obstetric histories from medical records at recruitment. Multivariable logistic regression analyses were performed to identify independent associations of induction and augmentation of labour with the clinical and non-clinical factors.

**RESULTS:** Among 9305 women, over two-fifth experienced labour induction (n=3936, 42.3%) and about a quarter had labour augmentation (n=2537, 27.3%). The majority who received labour induction/augmentation had at least one or more clinical indications, but around 34% did not have an indication. Compared with women with ≥2 indications, those with one (adjusted odds ratio 0.50, 95% confidence intervals 0.42 to 0.58) or no (0.24, 0.20 to 0.28) indication or with contraindications (0.12, 0.07 to 0.20) were less likely to be induced, adjusting for non-clinical characteristics. These associations were similar for augmentation of labour (0.71, 0.61 to 0.84, for one indication; 0.47, 0.39 to 0.55 for no indication; 0.17, 0.09 to 0.34 for contraindications). Several maternal demographic, healthcare utilization and socio-economic factors were independently associated with labour induction and augmentation.

**CONCLUSIONS:** Decisions about induction and augmentation of labour in our study population in India were largely guided by clinical recommendations but in nearly a third, there was no clinical indication based on guidelines. Further research is required to understand the complex influence of clinical need and socio-demographic factors on labour induction/augmentation in the context of risk and safety.

**What is already known on this topic:**

- Several established international and national guidelines recommend specific clinical indications and/or contraindications for induction of labour.
- Pregnant women are also given the option to decide on labour induction and/or augmentation after providing relevant information

**What this study adds:**

- Compared to women with at least two clinical indications as per guidelines, those with one or no indication or with contraindications were less likely to be induced or augmented, independent of other non-clinical maternal characteristics.
- In a third of the participants, there was no clinical indication for induction/augmentation of labour based on guidelines.
- Several non-clinical factors including maternal demographic, healthcare utilization and socio-economic factors influenced the decision for labour induction and augmentation, which in turn could be due to women’s choice or clinicians’ unconscious bias and warrants further research.

## Introduction

Globally, a maternity care crisis has been characterized by the soaring rate of caesarean sections^1^ but now also includes the rapidly growing use of induction and augmentation of labour. The labour induction and augmentation are usually recommended when the continuation of a pregnancy or prolongation of labour is judged to pose more of a clinical risk to maternal and/or fetal health than if the pregnancy were to end. The decision to induce or augment labour has to be balanced with its potential adverse maternal and neonatal outcomes.^2–6^ Several national and international clinical guidelines for labour induction have been established.^7–11^ While these guidelines identify both maternal and fetal clinical conditions as potential indications for induction/augmentation, there are variations in the specific indications and contraindications within them.^7^ ^11^ Some additionally suggest offering pregnant women the option to decide on labour induction/augmentation after providing them with relevant information.^7^ ^11^ This is consistent with the modern practice of shared decision making, enabling maternal autonomy through information provision and choice. Therefore, there may be clinical and non-clinical factors influencing the decisions for labour induction and augmentation.

Several studies have explored the influence of maternal and fetal risk factors on the decision to induce^2^ ^4^ ^12–17^ or augment labour.^18^ ^19^ These studies primarily focused on individual maternal indications such as hypertension and diabetes in pregnancy, with little consideration given to wider maternal and fetal factors for a more comprehensive understanding of the decision-making process. They also failed to account for the range of clinical opinion as to what are absolute and what are relative contraindications. Grade 3 and 4 placenta praevia are absolute contraindications to induction, whereas breech presentation and two or more previous caesarean sections would trigger a range of responses from clinicians. Moreover, previous studies analysed only a limited number of socio-demographic characteristics such as maternal age and education, which does not enable the exploration of how a wide range of socio-demographic factors might further influence decision-making for labour induction/augmentation. The majority of studies are also limited to Western countries (i.e. Belgium, Canada, United Kingdom, United States of America and Poland)^12–15^ ^17^ ^19^, while other studies were performed by continents (i.e. Latin America, Africa and Asia)^2^ ^4^ ^16^ rather than in individual countries except for one in Nepal.^18^

In India, maternal morbidity and mortality represents a major public health burden. Every year, nearly 30 million women become pregnant with 5 million facing life-threatening complications.^20^ More worryingly, India records over 45,000 maternal deaths annually, ranking it as the country with the second highest number of maternal deaths worldwide.^21^ The World Health Organization (WHO) Global Survey of 20 Indian facilities in 2007-2008 estimated that 12.8% of pregnant women had an induction of labour,^4^ but there is a lack of more contemporary data. The present study aimed to provide a comprehensive insight into induction and augmentation of labour in pregnant women in India through a multicentre large-scale prospective cohort study.^20^ Our specific objectives were to estimate the rates of labour induction and augmentation, and examine the influence of maternal and fetal clinical conditions, socio-demographic, lifestyle and healthcare utilization factors on the decision to induce and/or augment labour.

## Methods

### Study design and population

A prospective cohort study was conducted through the Maternal and perinatal Health Research collaboration, India (MaatHRI). The details for MaatHRI have been described elsewhere.^20^ In brief, MaatHRI is a hospital-based collaborative research platform established in September 2018 to undertake large-scale epidemiological research to improve maternal and perinatal health in a setting with a high burden of maternal mortality and morbidity. To date, this platform includes a network of 16 tertiary and community hospitals (11 Government and 5 private) across six states in India, namely Assam, Meghalaya, Chhattisgarh, Uttar Pradesh, Himachal Pradesh and Maharashtra.

In this cohort study, pregnant women were enrolled during their third trimester of pregnancy (≥28 weeks of gestation) between January 2018 and August 2023 from 13 of these hospitals. Women were eligible for inclusion if they were at least 18 years of age and planned a vaginal birth in the participating hospital. Those who planned an elective caesarean section were not eligible for this study. All recruited pregnant women were then followed-up during labour and childbirth and up to 48 hours postpartum.

### Baseline and follow-up assessments

At the recruitment visit, written informed consent was obtained and questionnaires were used by research nurses to collect information face-to-face from pregnant women about their own and their partner’s socio-demographic status and lifestyle factors. Information about the current pregnancy, obstetric and pre-existing medical histories were obtained from the medical records of participants. At the follow-up visit, labour and childbirth details were abstracted from medical records.

### Maternal and household characteristics

Information about both clinical and non-clinical factors^22^ were analysed in the study. These included maternal and fetal clinical conditions in the current pregnancy, maternal demographic and health characteristics (i.e. age at labour, parity, body mass index (BMI) in early pregnancy, gestational weight gain, previous pregnancy problems, pre-existing medical problems), healthcare utilization indicators (i.e. number of antenatal check-ups, duration of iron-folic acid (IFA) supplementation, healthcare professional managing childbirth), lifestyle factors (i.e. adverse lifestyles such as smoking, alcohol consumption, chewing betel nut or tobacco) and household socio-economic characteristics (i.e. religion, residence, living below poverty line (BPL), maternal education, partner’s occupation).

The maternal and fetal clinical conditions were classified into four groups based on guidelines for labour induction from the WHO^23^, the National Institute for Health and Care Excellence (NICE)^24^, the American College of Obstetricians and Gynecologists (ACOG)^25^, and the Federation of Obstetric and Gynecological Societies of India (FOGSI)^8–10^: (i) two or more clinical indications, (ii) one clinical indication, (iii) no clinical indications and (iv) contraindication/s for induction. This was used to generate a categorical variable ‘induction-related clinical indications’. The clinical indications are detailed in supplementary table 1. Contraindications to induction, accepting that there is some variation of opinion, were those with specific placental problems such as placenta praevia, ≥2 previous caesarean sections and fetal malpresentation including breech presentation (supplementary table 1), regardless of any aforementioned indication. Women with pre-existing or current pregnancy problems that were neither indications nor a contraindication, and those without any health issues were combined as “No indication” for purposes of the analysis.

Several factors recorded as continuous variables were categorised to facilitate interpretations, namely parity defined as number of completed pregnancies at ≥28 weeks (0, 1, 2-4, ≥5), number of antenatal check-ups (0-2, 3, 4, ≥5 visits) and duration of IFA supplementation (none, <100, 100-179, ≥180 days; where national initiative recommends supplementation for at least 180 days^26^). Adverse lifestyle factors were determined based on responses to questions on smoking status, tobacco consumption, chewing betel nut and alcohol consumption and classified into three categories: i) ‘never’ if all responses were ‘never’, ii) ‘past’ if any response was either ‘gave up prior to pregnancy’ or ‘gave up during pregnancy’ and iii) ‘current’ if any response was ‘current’.

Maternal weight in early pregnancy (centred at 10 weeks of gestation) and gestational weight gain per week were estimated from weight measures at the first antenatal visit and during the recruitment visit from medical records using mixed-effects linear regression models with random effects at hospital level^27^ and linear regressions at individual level, respectively. We then calculated maternal BMI in early pregnancy (to approximate pre-pregnancy BMI^28^) by dividing the estimated weight in kilograms (kg) at 10 weeks of gestation by height in meters squared (m^2^), and classified maternal BMI into four categories based on Asian cut-offs^29^: i) underweight (<18.5 kg/m^2^), ii) normal weight (18.5-22.9 kg/m^2^), iii) overweight (23-24.9 kg/m^2^) and iv) obese (≥25 kg/m^2^). For each BMI category in early pregnancy, the gestational weight gain per week was categorized as i) within, ii) below or iii) above the Institute of Medicine guidelines (underweight: 0.44–0.58 kg; normal weight: 0.35– 0.50 kg; overweight: 0.23–0.33 kg; obese: 0.17–0.27 kg).^30^

The partner’s (99% married; 1% single) occupation was classified into four levels based on the National Classification of Occupations 2015 in India^31^: i) unemployed, ii) partly skilled and unskilled, iii) skilled manual and non-manual and iv) professional, managerial and technical.

### Induction and augmentation of labour

Status concerning labour induction or augmentation were separately recorded in two binary variables (Yes or No); women who received both interventions are not mutually exclusive and were included in both variables. When induced labour or augmented labour was reported, further details about their indications and methods were noted. The methods of labour induction were categorised into three groups: i) mechanical only – if amniotomy, balloon catheter and/or membrane sweeping were conducted without any pharmacological agent, ii) pharmacological only – if misoprostol, oxytocin and/or other prostaglandins were used without any mechanical method, and iii) combination – if both mechanical and pharmacological methods were used. Similarly, the methods of labour augmentation were categorised into: i) mechanical only – if only amniotomy was conducted, ii) pharmacological only – if only oxytocin and/or other prostaglandins were used, and iii) combination – if both mechanical and pharmacological methods were used.

### Statistical analyses

Descriptive analyses were performed to summarise frequencies and proportions of labour induction and augmentations, their methods and clinician-reported indications. Maternal and household characteristics were compared between binary status of labour induction and augmentation, using chi-squared tests for categorical variables and t-tests for continuous variables.

To investigate the associations between clinical and non-clinical factors and labour induction or augmentation, three logistic regression models were computed for each outcome, with sequential addition of variables to the models based on a theoretical framework of the decision-making process for induction and augmentation (supplementary figure 1). Model 1 included only the ‘induction-related clinical indications’ variable which encompasses conditions that are usually the primary consideration for labour induction/augmentation. Model 2 additionally included maternal demographic, medical and obstetric characteristics. Model 3 expanded Model 2 by further adding healthcare utilization and lifestyle factors, and household socio-economic characteristics. A sensitivity analysis for labour augmentation was also conducted by including labour induction status in Model 3. All models were then compared using the area under the receiver operating characteristics curve (AUROC) analyses to assess the risk prediction of each model.

If an association between women’s socio-economic background and labour induction or augmentation was found, we further examined whether specific population groups were different by labour induction/augmentation as per clinical guidelines. We re-categorised the ‘induction-related clinical indications’ variable into three groups: i) no induction/augmentation ii) induction/augmentation with ≥1 clinical indication and iii) induction/augmentation with no-/contra-indications, and tested their associations with socio- economic factors using multivariable multinomial logistic regressions, adjusting for maternal demographic, medical and obstetric characteristics, healthcare utilization and lifestyle factors. For all regression analyses, missingness or the unknown category in most variables were treated as missing values given their low proportions (<6%). The unknown category for BPL status was treated as a separate group considering that it was reported by a high proportion (16.4%) of participants.

All statistical analyses were conducted using Stata 17.0 (StataCorp. 2021. Stata Statistical Software: Release 17. College Station, TX: StataCorp LLC) and all associations were considered to be significant at a two-tailed p value of <0.05.

## Results

### Study characteristics

A total of 9420 pregnant women were recruited, of whom 89 (∼1%) were lost to follow-up and one died before childbirth. After further excluding 25 women who later opted for elective caesarean sections, 9305 women were included in the analysis. More than 40% of these women had labour induced (n=3936, 42.3%), or in over a quarter of women labour was augmented (n=2537, 27.3%); with nearly one-fifth having both labour induction and augmentation (n=1789, 19.2%). The main clinician-recorded indications (not mutually exclusive) for induction of labour were fetal compromise (n=727, 18.5%), hypertensive disorders in pregnancy (n=640, 16.3%) and term pregnancy (n=498, 12.7%), and for labour augmentation were fetal compromise (n=788, 31.1%) and labour progression issues (n=602, 23.7%), see supplementary table 2. The most common methods used for labour induction or augmentation were pharmacological only (induction: n=2316, 58.8%; augmentation: n=1745, 68.8%), followed by mechanical only (induction: n=934, 23.8%; augmentation: n=555, 21.9%) and finally a combination (induction: n=686, 17.4%; augmentation: n=237, 9.3%).

### Factors associated with labour induction and augmentation

Table 1 shows the comparisons of maternal and household characteristics among included women with and without labour induction or augmentation. Most of the characteristics were significantly different between women who had their labour induced or augmented and those who did not. Among women with labour induction and/or augmentation (n=4684), about 33% (n=1560) had no clinical indication and 0.8% (n=35) had a contraindication. Within this ‘contraindication’ sub-group, the majority had malpresentation (n=20), followed by placenta problems (n=13) and 2-3 previous caesarean sections (n=2), and also a majority eventually had an emergency caesarean section (n=26, 74.3%). In general, most maternal and household characteristics were weakly correlated between each other (Spearman correlation, ρ <0.3), except for age and parity where the correlation was stronger (ρ = 0.443). The partner’s education level was not included in the analysis given its high correlation with women’s education level (ρ = 0.740). Also, both women’s marital status and occupation were not included due to the low proportions of being single (0.1%) and employed (1.6%).

**Table 1.** Characteristics of included participants with and without labour induction or augmentation in a prospective study in India.

| Maternal and household characteristics | No induction<br>N=5,369 | Induced labour<br>N=3,936 | P value | No augmentation<br>N=6,768 | Augmented labour<br>N=2,537 | P value |
| --- | --- | --- | --- | --- | --- | --- |
| Induction-related clinical indications (n, %) |  |  | <0.001 |  |  | <0.001 |
| ≥2 indications | 466 (8.7) | 914 (23.2) |  | 810 (12.0) | 570 (22.5) |  |
| 1 indication | 1,911 (35.6) | 1,824 (46.3) |  | 2,563 (37.9) | 1,172 (46.2) |  |
| No indication | 2,886 (53.8) | 1,171 (29.8) |  | 3,277 (48.4) | 780 (30.7) |  |
| Contraindications | 106 (2.0) | 27 (0.7) |  | 118 (1.7) | 15 (0.6) |  |
| Age at labour (mean (SD), years) | 24.85 (4.40) | 24.65 (4.50) | 0.033 | 24.73 (4.34) | 24.85 (4.72) | 0.26 |
| Parity (n, %) |  |  | <0.001 |  |  | <0.001 |
| 0 | 3,278 (61.1) | 2,963 (75.3) |  | 4,435 (65.5) | 1,806 (71.2) |  |
| 1 | 1,391 (25.9) | 572 (14.5) |  | 1,565 (23.1) | 398 (15.7) |  |
| 2-4 | 656 (12.2) | 373 (9.5) |  | 729 (10.8) | 300 (11.8) |  |
| ≥5 | 44 (0.8) | 28 (0.7) |  | 39 (0.6) | 33 (1.3) |  |
| BMI categories in early pregnancy (n, %) |  |  | <0.001 |  |  | <0.001 |
| Underweight (<18.5 kg/m<sup>2</sup>) | 1,564 (29.1) | 878 (22.3) |  | 1,698 (25.1) | 744 (29.3) |  |
| Normal weight (18.5-22.9 kg/m<sup>2</sup>) | 2,752 (51.3) | 2,349 (59.7) |  | 3,720 (55.0) | 1,381 (54.4) |  |
| Overweight (23-24.9 kg/m<sup>2</sup>) | 492 (9.2) | 364 (9.2) |  | 649 (9.6) | 207 (8.2) |  |
| Obesity (≥25 kg/m<sup>2</sup>) | 309 (5.8) | 221 (5.6) |  | 433 (6.4) | 97 (3.8) |  |
| missing | 252 (4.7) | 124 (3.2) |  | 268 (4.0) | 108 (4.3) |  |
| Gestational weight gain classifications (n, %) |  |  | <0.001 |  |  | <0.001 |
| Within guidelines | 754 (14.0) | 588 (14.9) |  | 988 (14.6) | 354 (14.0) |  |
| Below guidelines | 3,468 (64.6) | 2,752 (69.9) |  | 4,435 (65.5) | 1,785 (70.4) |  |
| Above guidelines | 749 (14.0) | 432 (11.0) |  | 910 (13.4) | 271 (10.7) |  |
| missing | 398 (7.4) | 164 (4.2) |  | 435 (6.4) | 127 (5.0) |  |
| Previous pregnancy problems (n, %) |  |  | 0.027 |  |  | 0.32 |
| No | 5,097 (94.9) | 3,717 (94.4) |  | 6,408 (94.7) | 2,406 (94.8) |  |
| Yes | 184 (3.4) | 170 (4.3) |  | 253 (3.7) | 101 (4.0) |  |
| Unknown | 88 (1.6) | 49 (1.2) |  | 107 (1.6) | 30 (1.2) |  |
| Pre-existing medical problems (n, %) |  |  | 0.21 |  |  | <0.001 |
| No | 5,254 (97.9) | 3,833 (97.4) |  | 6,649 (98.2) | 2,438 (96.1) |  |
| Yes | 114 (2.1) | 103 (2.6) |  | 118 (1.7) | 99 (3.9) |  |
| missing | 1 (0.0) | 0 (0.0) |  | 1 (0.0) | 0 (0.0) |  |
| Number of antenatal check-ups (n, %) |  |  | <0.001 |  |  | <0.001 |
| 0-2 | 1,428 (26.6) | 592 (15.0) |  | 1,580 (23.3) | 440 (17.3) |  |
| 3 | 1,396 (26.0) | 1,889 (48.0) |  | 1,884 (27.8) | 1,401 (55.2) |  |
| 4 | 1,451 (27.0) | 723 (18.4) |  | 1,776 (26.2) | 398 (15.7) |  |
| ≥5 | 1,094 (20.4) | 732 (18.6) |  | 1,528 (22.6) | 298 (11.7) |  |
| Duration of iron-folic acid supplementation (days) (n, %) |  |  | <0.001 |  |  | <0.001 |
| None | 144 (2.7) | 228 (5.8) |  | 183 (2.7) | 189 (7.4) |  |
| <100 | 1,750 (32.6) | 1,803 (45.8) |  | 2,053 (30.3) | 1,500 (59.1) |  |
| 100-179 | 2,839 (52.9) | 1,202 (30.5) |  | 3,439 (50.8) | 602 (23.7) |  |
| ≥180 | 636 (11.8) | 703 (17.9) |  | 1,093 (16.1) | 246 (9.7) |  |
| Healthcare professional managing childbirth (n, %) |  |  | <0.001 |  |  | <0.001 |
| Auxiliary nurse midwife/nurse | 2,839 (52.9) | 1,352 (34.3) |  | 3,088 (45.6) | 1,103 (43.5) |  |
| Resident doctor | 2,210 (41.2) | 2,365 (60.1) |  | 3,248 (48.0) | 1,327 (52.3) |  |
| Obstetrician | 121 (2.3) | 190 (4.8) |  | 216 (3.2) | 95 (3.7) |  |
| Others | 23 (0.4) | 3 (0.1) |  | 15 (0.2) | 11 (0.4) |  |
| missing | 176 (3.3) | 26 (0.7) |  | 201 (3.0) | 1 (0.0) |  |
| Adverse lifestyles (smoking, consumption of tobacco, betel nut or alcohol) (n, %) |  |  | <0.001 |  |  | <0.001 |
| Never | 3,617 (67.4) | 2,416 (61.4) |  | 4,543 (67.1) | 1,490 (58.7) |  |
| Past | 226 (4.2) | 121 (3.1) |  | 270 (4.0) | 77 (3.0) |  |
| Current | 1,526 (28.4) | 1,399 (35.5) |  | 1,955 (28.9) | 970 (38.2) |  |
| Religion (n, %) |  |  | <0.001 |  |  | <0.001 |
| Hindu | 4,271 (79.5) | 2,589 (65.8) |  | 5,330 (78.8) | 1,530 (60.3) |  |
| Muslim | 633 (11.8) | 1,154 (29.3) |  | 994 (14.7) | 793 (31.3) |  |
| Christian | 225 (4.2) | 155 (3.9) |  | 182 (2.7) | 198 (7.8) |  |
| Sikh/others | 240 (4.5) | 38 (1.0) |  | 262 (3.9) | 16 (0.6) |  |
| Residence (n, %) |  |  | <0.001 |  |  | <0.001 |
| Rural | 4,698 (87.5) | 3,254 (82.7) |  | 5,695 (84.1) | 2,257 (89.0) |  |
| Suburb/urban | 671 (12.5) | 682 (17.3) |  | 1,073 (15.9) | 280 (11.0) |  |
| Living below poverty line (BPL) (n, %) |  |  | <0.001 |  |  | <0.001 |
| BPL certificate/self-certified | 2,203 (41.0) | 2,457 (62.4) |  | 2,748 (40.6) | 1,912 (75.4) |  |
| not BPL | 2,074 (38.6) | 1,047 (26.6) |  | 2,720 (40.2) | 401 (15.8) |  |
| unknown | 1,092 (20.3) | 432 (11.0) |  | 1,300 (19.2) | 224 (8.8) |  |
| Woman's education (n, %) |  |  | <0.001 |  |  | <0.001 |
| Illiterate | 647 (12.1) | 270 (6.9) |  | 739 (10.9) | 178 (7.0) |  |
| ≤5th class | 729 (13.6) | 1,194 (30.3) |  | 1,085 (16.0) | 838 (33.0) |  |
| 6-12th class | 2,952 (55.0) | 1,953 (49.6) |  | 3,639 (53.8) | 1,266 (49.9) |  |
| ≥12th class | 1,017 (18.9) | 510 (13.0) |  | 1,273 (18.8) | 254 (10.0) |  |
| unknown | 24 (0.4) | 9 (0.2) |  | 32 (0.5) | 1 (0.0) |  |
| Husband's occupation (n, %) |  |  | <0.001 |  |  | <0.001 |
| Unemployed | 186 (3.5) | 402 (10.2) |  | 474 (7.0) | 114 (4.5) |  |
| Partly skilled and unskilled | 1,567 (29.2) | 1,215 (30.9) |  | 1,765 (26.1) | 1,017 (40.1) |  |
| Skilled manual and non-manual | 3,334 (62.1) | 2,138 (54.3) |  | 4,216 (62.3) | 1,256 (49.5) |  |
| Professional, managerial and technical | 275 (5.1) | 180 (4.6) |  | 306 (4.5) | 149 (5.9) |  |
| missing | 7 (0.1) | 1 (0.0) |  | 7 (0.1) | 1 (0.0) |  |

Figure 1 shows the associations of maternal and household characteristics with labour induction. Compared with women who had two or more induction-related clinical indications, those with one or no indication or with contraindication were significantly less likely to undergo induction of labour. These findings remained consistent after adjustment for other maternal and household characteristics in all three models (Model 3 - one indication: adjusted odds ratio (aOR): 0.50, 95% confidence interval (CI): 0.42, 0.58; no indication: aOR: 0.24, 95% CI: 0.20, 0.28; contraindication: aOR: 0.12, 95% CI: 0.07, 0.20). When simultaneously considering clinical indications, and maternal demographic and health characteristics (Model 2), primiparity and multiparity of 2-4 (vs. nulliparity), underweight (vs. normal weight) in early pregnancy and gestational weight gain above (vs. within) guidelines were less likely to be associated with induced labour. The odds of induction increased by 2% per year increase in age of the pregnant women at labour. The odds of induction was also higher for women with a gestational weight gain below the recommended guidelines and a history of previous pregnancy problems. With the exception of gestational weight gain, these associations remained significant even after adjusting for healthcare utilization, lifestyle, and household socio-economic factors in Model 3. Furthermore, women had a higher odds of induction if they had a higher number of antenatal check-ups (≥3 vs. 0- 2), if childbirth was managed by a resident doctor and obstetrician (vs. auxiliary nurse midwife/nurse) and if they did not receive any IFA supplementation (vs. ≥180 days of supplementation) during pregnancy. A lower level of education (≤5^th^ and 6-12^th^ vs. ≥12^th^ class), partner being unemployed (vs. in professional employment), BPL certified/self-certified (vs. not BPL), belonging to a minority religious Muslim or Christian background (vs. Hindu), and living in suburban/urban (vs. rural) areas were associated with increased odds of induction of labour. Overall, the AUROCs for predicting labour induction increased from 0.67 to 0.79 with the addition of non-clinical factors beyond clinical indications for induction (supplementary figure 2).

**Figure 1.**
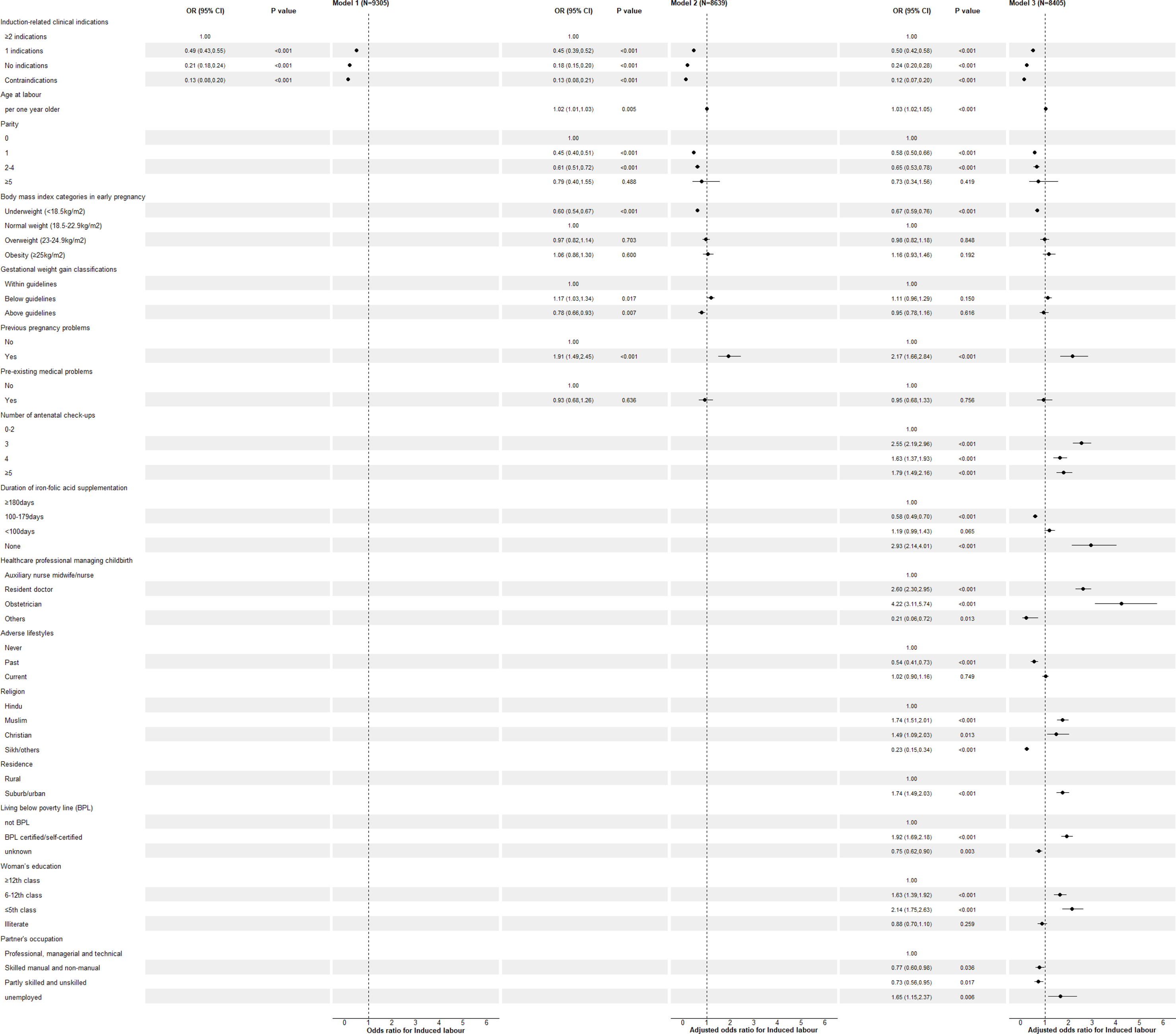
: Associations of maternal and household characteristics with labour induction in a prospective study in India

Figure 2 shows the associations of maternal and household characteristics with labour augmentation. The results were largely similar to those observed for induction of labour. Compared with women who had two or more induction-related clinical indications, those with one indication (aOR: 0.71, 95% CI: 0.61, 0.8 4), no indication (aOR: 0.47, 95% CI: 0.39, 0.55) and contraindications (aOR: 0.17, 95% CI: 0.09, 0.34) were slightly less likely to undergo labour augmentation, after adjustment for other maternal and household characteristics in all models. Also, per year increase in age at labour, being underweight in early pregnancy, having pre-existing medical problems, higher number of antenatal check-ups, lower duration of IFA supplementation, being managed by resident doctor, belonging to a minority religious Muslim or Christian background, BPL certified/self-certified and a having lower level of education were independently associated with higher odds of augmentation (Model 3). Upon adding labour induction status into Model 3, the observed associations mostly remained significant and women who had an induction of labour had more than three-fold higher odds of augmentation (supplementary figure 3). Overall, the AUROCs for predicting labour augmentation increased from 0.62 to 0.83 when maternal and household characteristics including healthcare utilization, lifestyle and socio-economic factors were added to the model, but remained unchanged when labour induction was additionally included (Supplementary figure 4).

**Figure 2:**
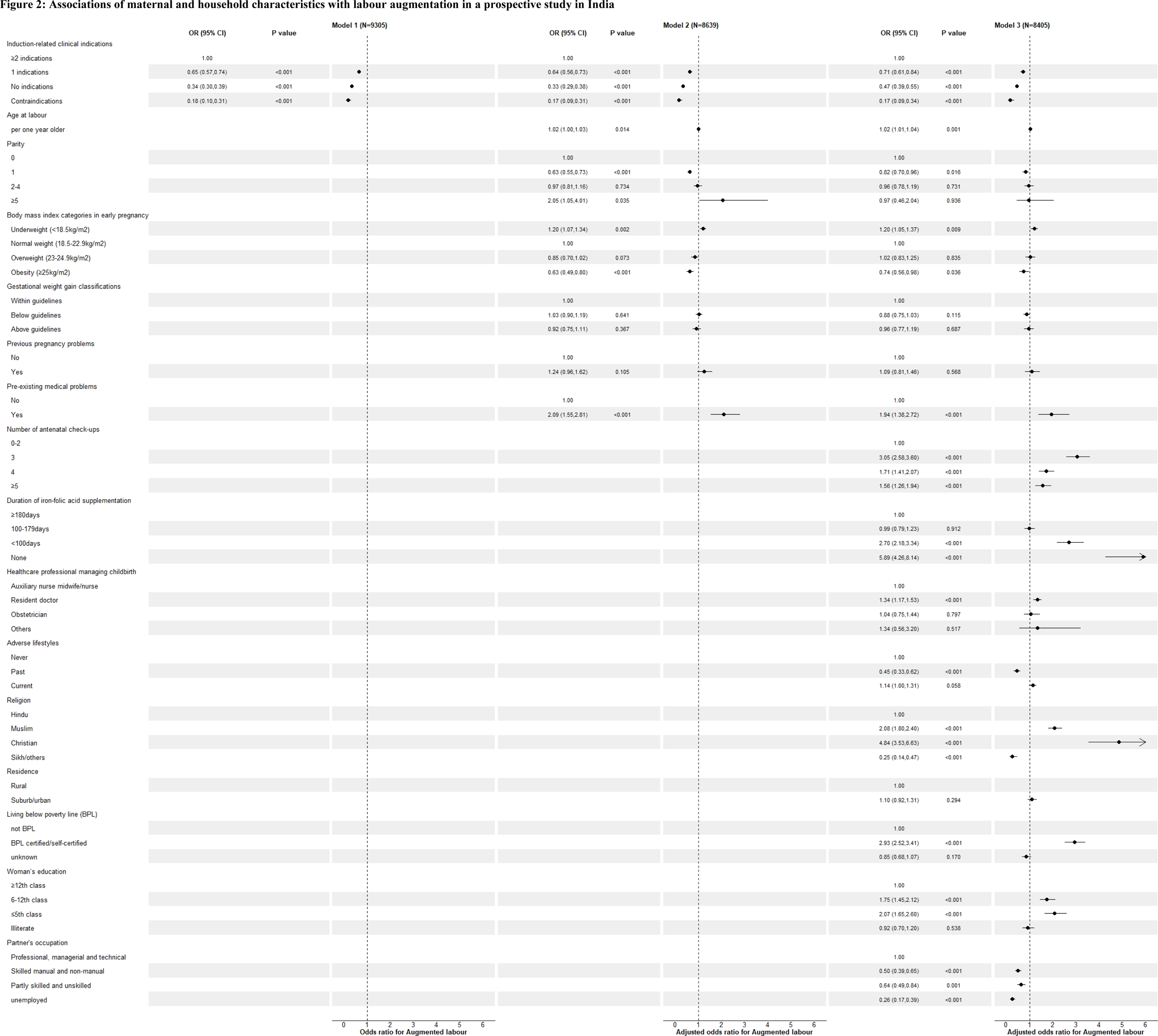
Associations of maternal and household characteristics with labour augmentation in a prospective study in India

Supplementary table 3 shows the adjusted associations of socio-economic factors with clinical indications for labour induction and augmentation. Compared with women with no induction or augmentation, women who belonged to minority religious Muslim or Christian background, lived in suburban or urban areas, reported BPL and had a lower level of education were more likely to have labour induction with both ≥1 clinical indications and/or with no-/contra-indication.

## Discussions

In this large prospective cohort study of pregnant women across 13 hospitals in India, a substantial rate of labour induction/augmentation was reported. Two out of every five women had labour artificially induced or over a quarter had their labour augmented; with nearly one-fifth having both induction and augmentation. Approximately two-thirds of the methods used for labour induction or augmentation were pharmacological only. Consistent with current guidelines for labour induction,^8–10^ ^23–25^ the greater the number of maternal and fetal clinical indications a woman had, the more likely her labour was to be induced, whereas women with any contraindication were less likely to be induced. Similarly, women with fewer clinical indications or contraindications for induction were less likely to undergo labour augmentation. Approximately 1% of women who were induced or augmented had clinical contraindications and approximately 33% did not have any documented indication. The associations with clinical indications were independent of other non-clinical maternal and household characteristics. In addition, maternal demographic characteristics (age at labour, parity and BMI in early pregnancy), healthcare utilization factors (number of antenatal check-ups, duration of IFA supplementation and healthcare professional managing childbirth) and socio-economic characteristics (religion, BPL status, maternal education and partner’s occupation) were independently associated with labour induction and augmentation. On further analyses, we found that women living in suburban/urban areas and from disadvantaged and minority socio-economic backgrounds were more likely to undergo induction and/or augmentation if they had clinical indications as well as if they had no indications or contraindications.

Compared with the WHO Global Survey of 20 Indian healthcare facilities in 2007-2008,^4^ the present findings suggest a potential threefold increase in the prevalence of labour induction in India (from 12.8% in the WHO survey to 42.3% in our study) over two decades, with induction using pharmacological agents alone being the predominant method (77.4% in the WHO survey vs. 58.8% in our study). Also, the WHO Global Survey^4^ and our study consistently found that fetal-related complications and hypertensive disorders in pregnancy were the most common indications for induction (representing at least 10% of inductions), and identified similar proportions of induction of labour without any clinical indication as per guidelines^8–10^ ^23–25^ (32.1% in WHO survey vs. ∼31% in our study). The increased utilization of induction of labour may be partially explained by improved diagnosis or increasing prevalence of maternal and/or fetal complications, but this also depends on what is reported in the labour management records. We added that labour was generally augmented using pharmacological agents only, and the reported indications were not solely conditions affecting the progress of labour, but also pregnancy complications especially poor fetal condition, although the two indications are related as poor progress in labour could lead to poor fetal condition.

The current study largely expands previous analyses in other populations,^2^ ^4^ ^12–19^ encompassing a comprehensive range of both maternal and fetal clinical factors in relation to labour induction and augmentation. Most studies have individually examined health complications in mothers and/or fetus, and reported associations of various clinical conditions, including hypertensive disorders in pregnancy and non-indications^8^ ^10^ such as anxiety, depression and urinary tract infection, with labour induction^2^ ^4^ ^12^ ^13^ or augmentation^18,19^. One of these studies also showed several contraindications (i.e. placental abruption, placenta praevia, cord prolapse and breech or malpresentation) to be less likely to be associated with labour induction.^12^ We considered and classified all recorded maternal and fetal clinical conditions as per guidelines for labour induction^8–10^ ^23–25^ and showed that fewer indications and any contraindications were less likely to be associated with labour induction and augmentation in the Indian study population, even after adjusting for maternal healthcare utilization, lifestyle and socio-demographic characteristics, thus highlighting generally good clinical practice among our study healthcare facilities. A very small number of women with contraindications were however induced (n=27) or augmented (n=15), where the contraindications were identified during antenatal visits or recorded as indications for emergency caesarean section, suggesting that clinicians may have potentially missed or not fully considered all pertinent information in medical records, or not adequately assessed women’s and their fetus’ health conditions before labour intervention. In contrast, a study in the United Kingdom that classified pregnant women into four categories (i.e. no pregnancy complications, pregnancy complications not usually associated with labour induction, pregnancy complications associated with labour induction, and others) observed all four groups to be associated with with labour induction.^17^

Through the analyses of a wide range of non-clinical factors, this study highlighted that several maternal demographic, healthcare utilization and socio-economic factors could influence the decision to induce and/or augment labour. The combination of these non-clinical factors increased the predictions for labour induction and augmentation, as noted by the increased AUROCs explained by the models including non-clinical factors. Similar to our findings, previous studies in the United States of America, United Kingdom, Belgium, and Latin American, African and Asian regions have reported that pregnant women who were in the older age groups,^2^ ^16^ had lower education level,^14^ ^16^ ^17^ more antenatal visits/prenatal care^4, 12^ ^16^ or childbirth attended by physicians,^12^ or lived in urban areas^16^ tended to undergo induction of labour. Associations with labour augmentation were however mostly different in previous studies, with higher odds for pregnant women who were younger or nulliparous, or had a higher education level in Nepal^18^ or had a higher BMI in Poland^19^.

Our findings of independent associations for clinical indications, as well as healthcare utilization and socio-demographic factors indicate a complex involvement of clinical practices and maternal characteristics, beyond pathologies alone, in influencing the decisions for inducing and augmenting labour. It is possible that clinicians lower their threshold for labour interventions based on other risk factors due to various reasons, irrespective of clinical indications. Older pregnant women or women with previous pregnancy problems or pre-existing medical problems, from lower socio-economic status (proxied by lower education level and BPL certified/self-certified) or from religious minority backgrounds (such as Muslim and Christian) might be thought to be at a higher risk of adverse maternal and fetal outcomes^32^ and hence were more likely to be offered labour induction and/or augmentation. Indeed on further analyses, women from disadvantaged or minority backgrounds were found to undergo induction/augmentation if they had one or more clinical indications, but they also had a higher odds of having induction/augmentation if they had no indication or a contraindication. This suggests that these groups of women may be also considered as the disadvantaged and vulnerable minorities within society, prompting clinicians to favour labour interventions to minimize any potential (despite lower) risk of prolonged pregnancy- or labour-related complications; such a tendency may arise from pressure by public or institutional expectations, or fear of potential abuse or litigation.^32^ Similar pressure or fear may also lead resident doctors or obstetricians who manage childbirth care to opt for labour interventions aiming to facilitate safe labour and childbirth.^32^ Alternatively, clinicians may be guided by the changes in societal norms with a greater emphasis on shared decision making and a woman’s right to choose, which unintentionally allows certain groups of women leaning towards labour induction/augmentation. Women who came from lower socio- economic status or lived in suburb or urban areas, or belonged to religious minorities may tend to view induction/augmentation as a better option (such as for pain relief) due to poor access to information from unreliable sources such as social media, or based on cultural or religious beliefs.^32^ Again, it is possible that women who had more antenatal visits possibly had higher expectations of labour care including labour interventions.

This is the first large-scale prospective cohort study in India that comprehensively examined the clinical and non-clinical factors influencing decisions for labour intervention. However, we examined only 13 hospitals in India, which limits the generalizability of our findings to all healthcare facilities in India and other settings. Systematic bias may exist if clinicians tended to selectively report relevant clinical indications for women undergoing induced or augmented labour, but we carefully minimized this bias by taking into account all maternal and fetal clinical conditions, which were prospectively ascertained at recruitment and after childbirth from medical records, instead of relying only on clinician-reported indications. The small number of women who had a placental problem but were still induced or augmented might have been misclassified as contraindicated due to insufficient information. These could have been women with minor grade placenta praevia but as information on grade of severity was not available, women with any reported placental problems were classified under the contraindication group. Furthermore, contraindications for induction are controversial and mainly provided by the FOGSI but not others.^8–10^ ^23–25^ Moreover, socio-economic characteristics and lifestyles may be subject to reporting bias for various reasons such as social desirability, although these are likely to be nondifferential misclassifications which bias associations towards the null. The status of labour induction and augmentation may have been cross-misclassified as certain methods used to induce or augment labour overlap such as amniotomy^17^ and oxytocin.^33^ However, we have minimized the potential biases by ensuring no duplications in the reported methods of labour induction and augmentation for each woman. Although our analyses included a wide range of maternal healthcare utilization, lifestyle and socio-demographic characteristics, our observed associations may be affected by other unmeasured risk factors such as age at first pregnancy and distance to the health facility. We did not include labour duration in the augmentation analysis models to avoid overadjustment as labour duration could be an outcome of the augmentation process itself. Finally, we cannot rule out the possibility of chance findings as we tested the associations of induction and augmentation with a wide range of non-clinical factors.

In conclusion, our analyses of pregnant women across multiple hospitals in India identified high proportions of women undergoing labour induction and/or augmentation, predominantly using pharmacological agents. We found that pregnant women with fewer indications and those with contraindications, as per guidelines for labour induction, were less likely to be induced or augmented, suggesting good clinical practice in general, although we cannot ignore that some women (a third) with no apparent indication or with contraindications were induced and/or augmented. Our findings importantly highlight that non-clinical factors such as maternal healthcare utilization and socio-demographic characteristics, beyond the commonly considered maternal and fetal complications, could potentially influence the decision-making process for labour interventions in India, raising concern about clinical practices that lack a robust evidence base. Future in-depth research is needed to understand the complex influence of clinical need and socio-demographic factors on labour induction/augmentation in the context of risk and safety to enable women to be properly informed about the consequences of labour induction and augmentation. Replicating similar studies in other settings is crucial to determine the extent of non-clinical influences on labour induction and augmentation, which in turn can contribute towards improving local clinical guidelines, empowering women with adequate and appropriate information for decision-making and mitigating clinicians’ unconscious bias, amidst the widespread utilization of the invasive labour induction/augmentation.

## Contributors

TSC and MN contributed to the study conceptualisation. MN designed the study and developed the methodology. TSC conducted the statistical analysis and wrote the first draft of paper. TSC, CO, DC, SSC and MN interpreted the findings. FZ, CSV, AV, SR, SSC, GD, PM, SK, RM, SC, AR, AB, IR, BM and OKB are collaborators and investigators for the study, and contributed equally to developing the study and led the work in their respective institution. RD is MaatHRI project manager and supervised data collection, entry and compilation. MK and JK are advisors and contributed to developing the study. All authors read and approved the final version of the manuscript. The corresponding author attests that all listed authors meet authorship criteria and that no others meeting the criteria have been omitted.

## Competing interests

All authors have completed the ICMJE uniform disclosure form at www.icmje.org/disclosure-of-interest/ and declare: support from Medical Research Council Career Development and Transition Support Award for the submitted work; no financial relationships with any organisations that might have an interest in the submitted work in the previous three years; no other relationships or activities that could appear to have influenced the submitted work.

The corresponding author (MN) affirms that the manuscript is an honest, accurate, and transparent account of the study being reported; that no important aspects of the study have been omitted; and that any discrepancies from the study as planned (and, if relevant, registered) have been explained.

Dissemination to participants and related patient and public communities: MaatHRI has a community engagement and involvement group who are actively involved in all aspects of the studies including dissemination. Participants and public will be notified of the results through their clinicians, our newsletters, and the MaatHRI website.

Provenance and peer review: Not commissioned; externally peer reviewed.

## Ethics approval

This cohort study was performed in accordance with the principles of the Declaration of Helsinki. All participants provided their written informed consent. Ethics approvals were obtained from the institutional review boards of each coordinating Indian institution, the Government of India’s Health Ministry’s Screening Committee, the Indian Council of Medical Research, New Delhi and the Oxford Tropical Research Ethics Committee (OxTREC), University of Oxford, UK.

## Data sharing

Data are available upon reasonable request by contacting the corresponding author.

## Data Availability

Data are available upon reasonable request by contacting the corresponding author.

