## supplementary for "Risk factors for labour induction and augmentation: a multicentre prospective cohort study in India"

Tuck Seng Cheng, postdoctoral researcher[1], Farzana Zahir, associate professor[2], Carolin Solomi V, consultant[3], Ashok Verma, professor[4], Sereesha Rao, assistant professor[5], Saswati Sanyal Choudhury, professor[6], Gitanjali Deka, professor[7], Pranabika Mahanta, assistant professor[8], Swapna Kakoty, associate professor[9], Robin Medhi, professor[9], Shakuntala Chhabra, professor[10], Anjali Rani, associate professor[11], Amrit Bora, senior medical officer[12], Indrani Roy, consultant[13], Bina Minz, consultant[14], Omesh Kumar Bharti, state epidemiologist[15], Rupanjali Deka, project manager[16], Charles Opondo, associate professor[17], David Churchill, professor[18,19], Marian Knight, professor[1], Jennifer J Kurinczuk, emeritus professor[1], Manisha Nair, associate professor[1]

[1]National Perinatal Epidemiology Unit, Nuffield Department of Population Health, Oxford University, Oxford, UK

[2]Department of Obstetrics and Gynaecology, Assam Medical College, Dibrugarh, Assam, India

[3]Department of Obstetrics and Gynaecology, Makunda Christian Leprosy and General Hospital, Karimganj, Assam, India

[4]Department of Obstetrics and Gynaecology, Dr Rajendra Prasad, Government Medical College, Kangra, Tanda, Himachal Pradesh, India

[5]Department of Obstetrics and Gynaecology, Silchar Medical College and Hospital, Silchar, Assam, India

[6]Department of Obstetrics and Gynaecology, Gauhati Medical College and Hospital, Guwahati, Assam, India

[7]Department of Obstetrics and Gynaecology, Tezpur Medical College, Tezpur, India

[8]Department of Obstetrics and Gynaecology, Jorhat Medical College and Hospital, Jorhat, Assam, India

[9]Department of Obstetrics and Gynaecology, Fakhruddin Ali Ahmed Medical College and Hospital, Barpeta, Assam, India

[10]Department of Obstetrics and Gynaecology, Mahatma Gandhi Institute of Medical Sciences, Sevagram, Maharashtra, India

[11]Department of Obstetrics and Gynaecology, Banaras Hindu University Institute of Medical Sciences, Varanasi, Uttar Pradesh, India

[12]Department of Obstetrics and Gynaecology, Sonapur District Hospital, Assam, India.

[13]Department of Obstetrics and Gynaecology, Nazareth Hospital, Shillong, Meghalaya, India

[14]Department of Obstetrics and Gynaecology, Sewa Bhawan Hospital Society, Chattisgarh, India

[15]State Institute of Health and Family Welfare, Department of Health & Family Welfare, Government of Himachal Pradesh, India

[16]MaatHRI Project, Srimanta Sankaradeva University of Health Sciences, Guwahati, Assam, India

[17]Department of Medical Statistics, London School of Hygiene & Tropical Medicine, London, UK

[18]Department of Obstetrics and Gynaecology, The Royal Wolverhampton NHS Trust, UK

[19]Research Institute for Healthcare Science, University of Wolverhampton, UK

Corresponding author

Associate Professor Manisha Nair

National Perinatal Epidemiology Unit, Nuffield Department of Population Health, Oxford University, Old Road Campus, Headington, Oxford OX3 7LF, UK

**Supplementary table 1: Maternal and fetal indications and contraindications for induction of labour according to guidelines***

| Indications | Contraindications |
|---|---|
| • Pre-existing or current pregnancy health problems<br> ▪ Pre-existing diabetes mellitus or gestational diabetes mellitus<br> ▪ Hypertension disorders in pregnancy including pre-existing hypertension, pregnancy-induced hypertension, preeclampsia and eclampsia<br> ▪ Antepartum haemorrhage<br> ▪ Premature rupture of membrane<br> ▪ Chorioamnionitis<br> ▪ Oligohydramnios<br> ▪ Obstetric cholestasis<br> ▪ Isoimmunisation<br> ▪ Renal disease<br> ▪ Cardiac problem<br>• Current fetal health problems<br> ▪ Structural defects<br> ▪ Intrauterine demise<br> ▪ Small for gestational age (based on birthweight <2500g)<br>• Post-term pregnancy (≥41 weeks of gestation calculated from last menstrual period)<br>• Multiple gestation | • Specific placental problems such as a major placenta praevia<br>• ≥2 previous caesarean sections<br>• Fetal malpresentation including breech presentation |

*the World Health Organization[23], the National Institute for Health and Care Excellence[24], the American College of Obstetricians and Gynecologists[25], and the Federation of Obstetric and Gynecological Societies of India[8,10,26]

**Supplementary figure 1: Theoretical framework of the decision-making process for labour induction and augmentation**

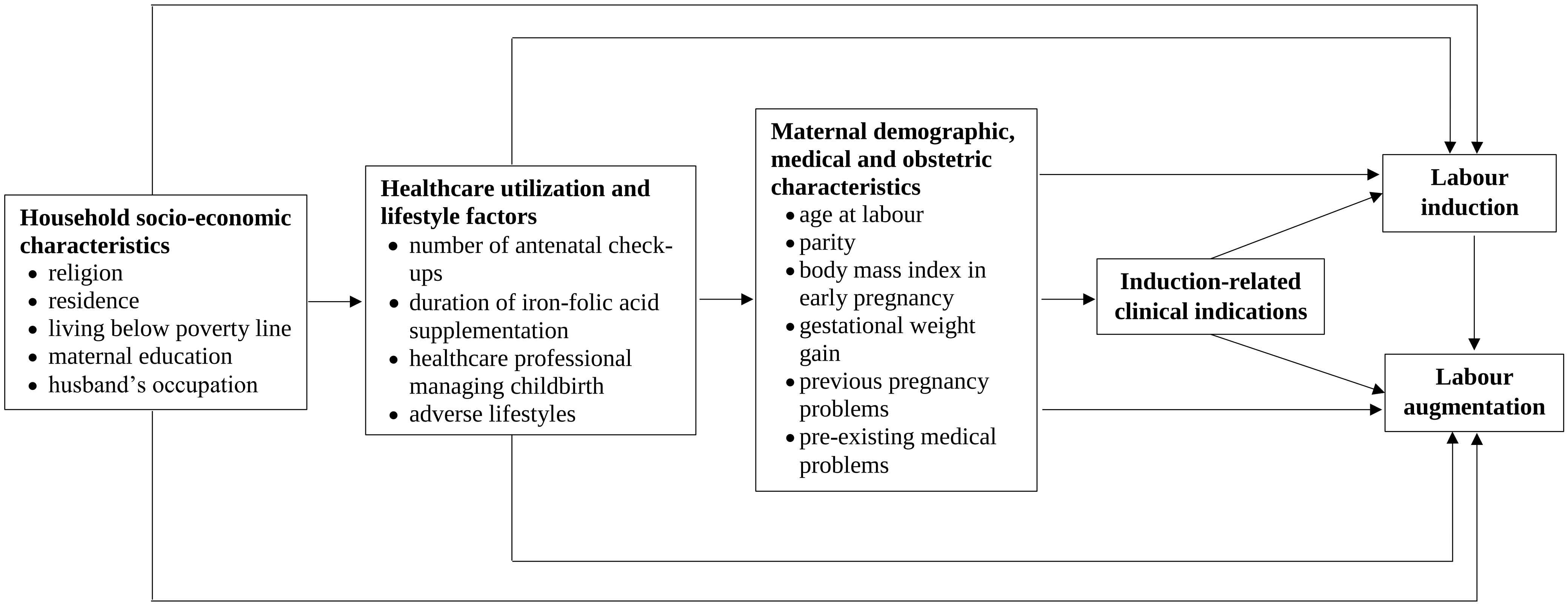

**Supplementary table 2: Frequencies of clinician-reported indications for labour induction and augmentation in the study hospitals in India**

| Reported indications* | Labour induction | | Labour augmentation | |
|---|---|---|---|---|
| | N | % | N | % |
| *Maternal factors* | | | | |
| Pregnancy-induced hypertension/ hypertension | 588 | 14.94 | 28 | 1.10 |
| Term pregnancy | 498 | 12.65 | 222 | 8.75 |
| Post-term pregnancy | 363 | 9.22 | 47 | 1.85 |
| Premature rupture of membrane | 340 | 8.64 | 96 | 3.78 |
| Oligohydramnios | 184 | 4.67 | 6 | 0.24 |
| Rupture of membrane | 134 | 3.40 | 48 | 1.89 |
| Preterm pregnancy | 114 | 2.90 | 36 | 1.42 |
| Early labour | 51 | 1.30 | 3 | 0.12 |
| Labour pain | 48 | 1.22 | 26 | 1.02 |
| Pre-eclampsia | 44 | 1.12 | 7 | 0.28 |
| Anaemia | 42 | 1.07 | 12 | 0.47 |
| Latent labour | 34 | 0.86 | 4 | 0.16 |
| No labour | 26 | 0.66 | 10 | 0.39 |
| Cholestasis | 16 | 0.41 | - | - |
| Gestational diabetes mellitus/ diabetes | 13 | 0.33 | 2 | 0.08 |
| Previous caesarean sections | 13 | 0.33 | - | - |
| Anhydraminious | 12 | 0.30 | - | - |
| Antepartum haemorrhage | 10 | 0.25 | 1 | 0.04 |
| Eclampsia | 8 | 0.20 | 4 | 0.16 |
| Grand multiparity | 8 | 0.20 | - | - |
| Rhesus negative | 7 | 0.18 | - | - |
| Short stature | 6 | 0.15 | - | - |
| Hypothyroidism | 6 | 0.15 | 1 | 0.04 |
| Polyhydraminious | 5 | 0.13 | - | - |
| Fetal demise in previous pregnancy | 5 | 0.13 | - | - |
| Elderly primigravida | 4 | 0.10 | - | - |
| Multiple gestation | 4 | 0.10 | 1 | 0.04 |
| Oedema | 4 | 0.10 | - | - |
| Meconium stained liquor | 4 | 0.10 | 14 | 0.55 |
| Poor obstetric history | 4 | 0.10 | - | - |
| Haemoglobin E disease | 3 | 0.08 | 2 | 0.08 |
| Uneffaced cervix | 3 | 0.08 | 5 | 0.20 |
| Deteriorating maternal condition | 2 | 0.05 | - | - |
| Chorioamnionitis | 2 | 0.05 | 1 | 0.04 |
| Infertility | 2 | 0.05 | - | - |
| Other maternal factors† | 13 | 0.33 | - | - |
| *Fetal factors* | | | | |
| Fetal compromise | 727 | 18.47 | 788 | 31.06 |
| Decreased fetal movement | 127 | 3.23 | 3 | 0.12 |
| Low birth weight / IUGR | 46 | 1.17 | 6 | 0.24 |
| Fetal demise | 26 | 0.66 | 17 | 0.67 |
| Fetal distress | 24 | 0.61 | - | - |
| Breech presentation | 8 | 0.20 | - | - |
| Non-reassuring fetal heat rate | 6 | 0.15 | 3 | 0.12 |

| | | | | |
|---|---|---|---|---|
| Abnormal Doppler | 4 | 0.10 | - | - |
| Mobile fetal head | 4 | 0.10 | 1 | 0.04 |
| Big baby | 2 | 0.05 | 1 | 0.04 |
| Other fetal factors‡ | 2 | 0.05 | 1 | 0.04 |
| *Labour-related factors* | | | | |
| Prolonged labour / no or slow progress in labour¥ | 116 | 2.95 | 203 | 8.00 |
| To accelerate labour¥ | 17 | 0.43 | 223 | 8.79 |
| Reported induction failure¥ | 11 | 0.28 | 10 | 0.39 |
| Poor or inadequate uterine contraction | - | - | 495 | 19.51 |

*not mutually exclusive (i.e. women could have >1 indication)

†including cardiac problems, renal problems, placenta problems, abortion, history of abortion, ascites, breathing difficulty, difficulty in childbirth in previous pregnancies, abnormal liver function test, obesity, history of preterm childbirth, high risk pregnancy

‡including birth asphyxia, cephalopelvic disproportion

%, percentages calculated by dividing the number of a particular reported indication separately by total induced (n=3936) or augmented (n=2537) labour

¥Note: These indicate that the woman was already in labour, but as these were mentioned as reasons for induction of labour, we have reported the information as such.

**Supplementary figure 2: Area under the receiver operating characteristics (AUROC) curve for prediction models for labour induction**

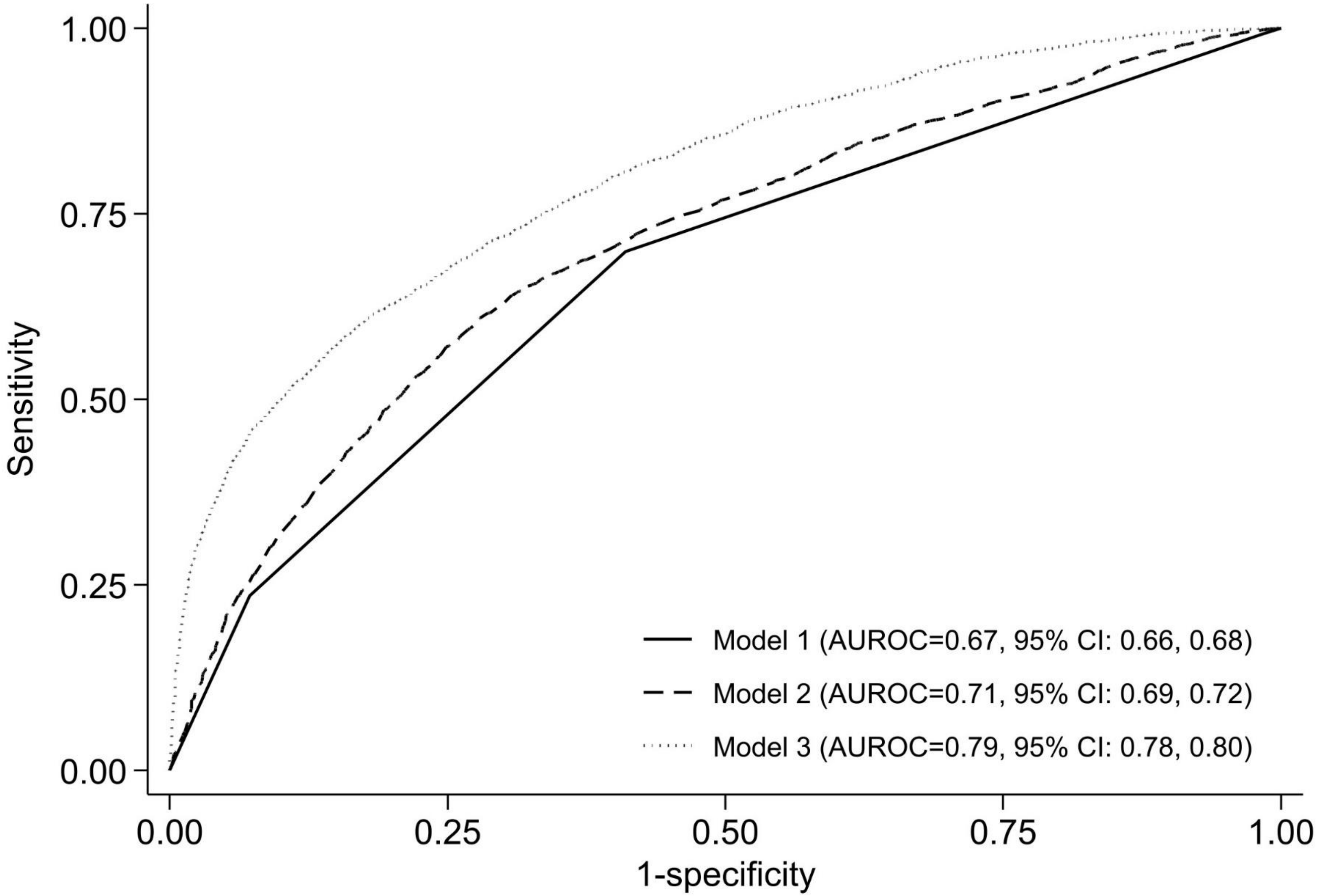

**Supplementary figure 3: Associations of maternal and household characteristics with labour augmentation, additionally adjusted for labour induction, in a prospective study in India**

| | OR (95% CI) | P value |
|---|---|---|
| Induced labour | | |
| No | 1.00 | |
| Yes | 3.35 (2.93,3.82) | <0.001 |
| Induction-related clinical indications | | |
| ≥2 indications | 1.00 | |
| 1 indications | 0.85 (0.72,1.00) | 0.053 |
| No indications | 0.66 (0.55,0.78) | <0.001 |
| Contraindications | 0.29 (0.15,0.59) | <0.001 |
| Age at labour | | |
| per one year older | 1.02 (1.00,1.03) | 0.021 |
| Parity | | |
| 0 | 1.00 | |
| 1 | 0.93 (0.79,1.09) | 0.369 |
| 2-4 | 1.07 (0.87,1.32) | 0.523 |
| ≥5 | 1.01 (0.48,2.15) | 0.969 |
| Body mass index categories in early pregnancy | | |
| Underweight (<18.5kg/m2) | 1.32 (1.15,1.52) | <0.001 |
| Normal weight (18.5-22.9kg/m2) | 1.00 | |
| Overweight (23-24.9kg/m2) | 1.04 (0.84,1.28) | 0.741 |
| Obesity (≥25kg/m2) | 0.70 (0.53,0.92) | 0.012 |
| Gestational weight gain classifications | | |
| Within guidelines | 1.00 | |
| Below guidelines | 0.85 (0.72,1.01) | 0.059 |
| Above guidelines | 0.97 (0.78,1.21) | 0.770 |
| Previous pregnancy problems | | |
| No | 1.00 | |
| Yes | 0.89 (0.66,1.21) | 0.463 |
| Pre-existing medical problems | | |
| No | 1.00 | |
| Yes | 2.04 (1.43,2.89) | <0.001 |
| Number of antenatal check-ups | | |
| 0-2 | 1.00 | |
| 3 | 2.51 (2.12,2.98) | <0.001 |
| 4 | 1.47 (1.21,1.79) | <0.001 |
| ≥5 | 1.35 (1.08,1.68) | 0.008 |
| Duration of iron-folic acid supplementation | | |
| ≥180days | 1.00 | |
| 100-179days | 1.18 (0.94,1.49) | 0.143 |
| <100days | 2.84 (2.28,3.55) | <0.001 |
| None | 5.11 (3.67,7.11) | <0.001 |
| Healthcare professional managing childbirth | | |
| Auxiliary nurse midwife/nurse | 1.00 | |
| Resident doctor | 1.06 (0.92,1.22) | 0.423 |
| Obstetrician | 0.78 (0.56,1.08) | 0.132 |
| Others | 1.96 (0.82,4.64) | 0.128 |
| Adverse lifestyles | | |
| Never | 1.00 | |
| Past | 0.50 (0.36,0.70) | <0.001 |
| Current | 1.17 (1.02,1.35) | 0.026 |
| Religion | | |
| Hindu | 1.00 | |
| Muslim | 1.90 (1.64,2.20) | <0.001 |
| Christian | 4.64 (3.36,6.41) | <0.001 |
| Sikh/others | 0.38 (0.21,0.71) | 0.002 |
| Residence | | |
| Rural | 1.00 | |
| Suburb/urban | 1.00 (0.83,1.20) | 0.983 |
| Living below poverty line (BPL) | | |
| not BPL | 1.00 | |
| BPL certified/self-certified | 2.57 (2.20,3.00) | <0.001 |
| unknown | 0.88 (0.70,1.11) | 0.298 |
| Woman's education | | |
| ≥12th class | 1.00 | |
| 6-12th class | 1.64 (1.34,1.99) | <0.001 |
| ≤5th class | 1.84 (1.46,2.32) | <0.001 |
| Illiterate | 0.95 (0.72,1.24) | 0.689 |
| Partner's occupation | | |
| Professional, managerial and technical | 1.00 | |
| Skilled manual and non-manual | 0.52 (0.40,0.68) | <0.001 |
| Partly skilled and unskilled | 0.68 (0.52,0.90) | 0.007 |
| unemployed | 0.23 (0.15,0.34) | <0.001 |

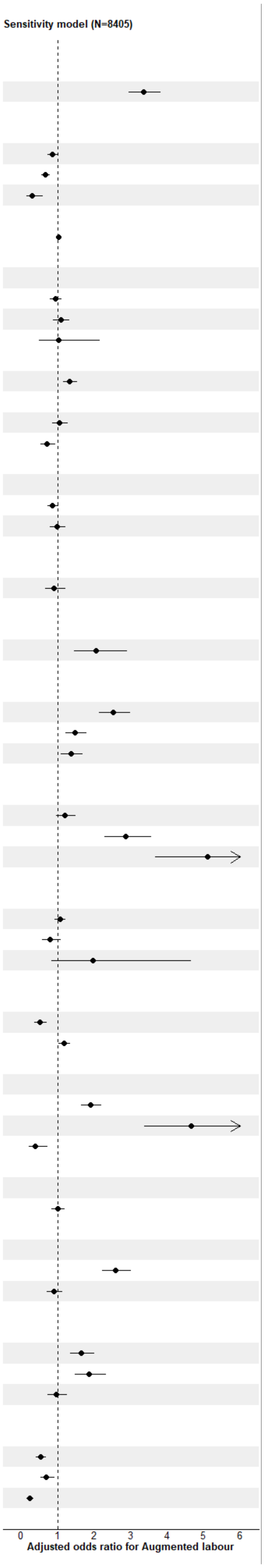

**Supplementary figure 4: Area under the receiver operating characteristics (AUROC) curve for prediction models for labour augmentation**

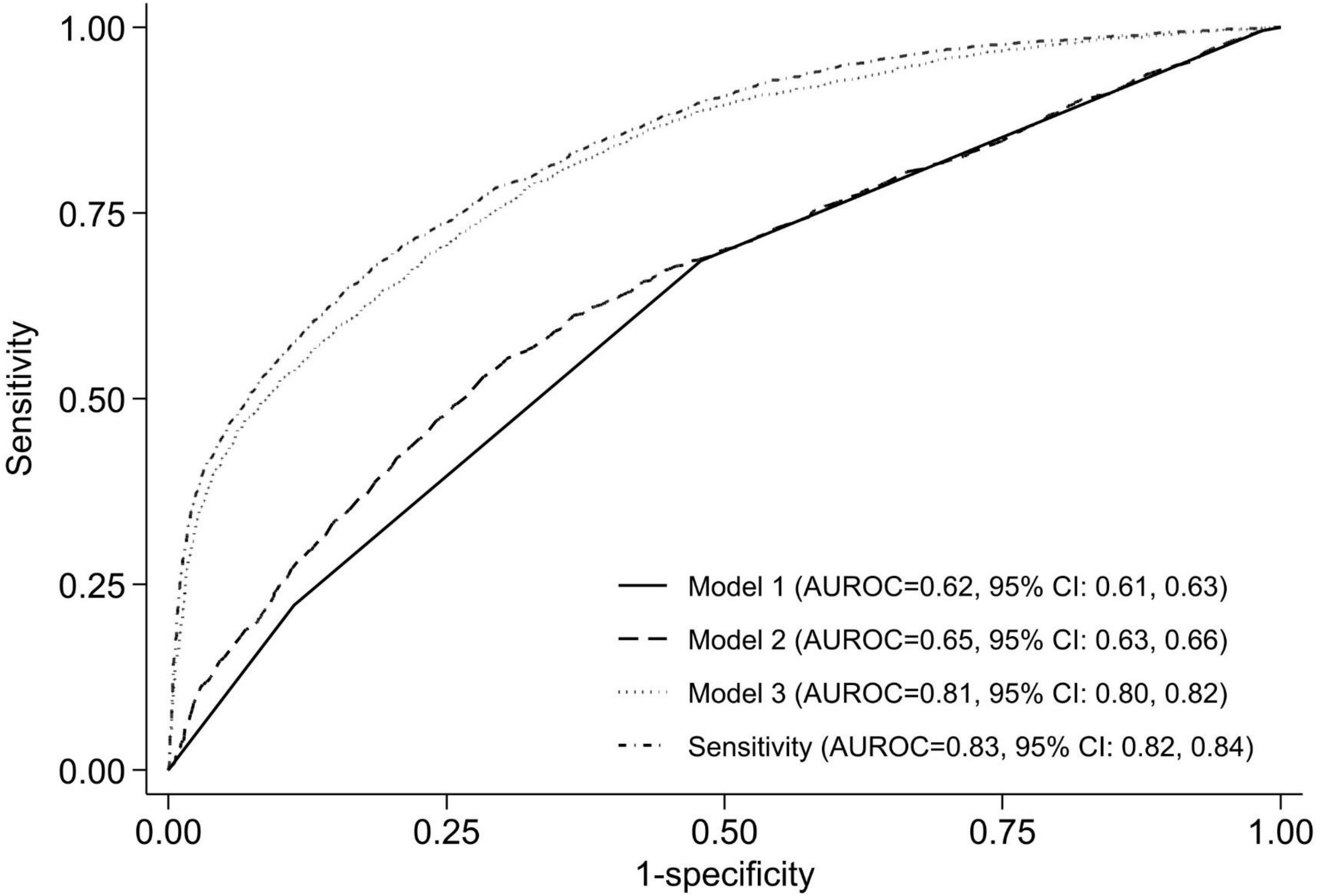

**Supplementary table 3: Adjusted associations of socio-economic factors with clinical indications for labour induction and augmentation in a prospective study in India‡**

| | Labour induction (n=8405) | | Labour augmentation (n=8405) | |
|---|---|---|---|---|
| | **Adjusted OR (95% CI)** | **P value** | **Adjusted OR (95% CI)** | **P value** |
| **No induction/augmentation** | No induction (base outcome) | | No augmentation (base outcome) | |
| **Induction/augmentation with ≥1 clinical indications** | | | | |
| Religion | | | | |
| Hindu | 1.00 | | 1.00 | |
| Muslim | 2.30 (1.98, 2.67) | <0.001 | 2.45 (2.09, 2.88) | <0.001 |
| Christian | 1.66 (1.16, 2.37) | 0.005 | 4.51 (3.09, 6.57) | <0.001 |
| Sikh/others | 0.26 (0.16, 0.43) | <0.001 | 0.16 (0.06, 0.43) | <0.001 |
| Residence | | | | |
| Rural | 1.00 | | 1.00 | |
| Suburb/urban | 1.64 (1.38, 1.95) | <0.001 | 0.98 (0.79, 1.22) | 0.857 |
| Living below poverty line (BPL) | | | | |
| not BPL | 1.00 | | 1.00 | |
| BPL certificate/self-certified | 2.20 (1.90, 2.55) | <0.001 | 3.19 (2.65, 3.85) | <0.001 |
| unknown | 0.99 (0.80, 1.22) | 0.932 | 1.14 (0.87, 1.49) | 0.336 |
| Woman's education | | | | |
| ≥12th class | 1.00 | | 1.00 | |
| 6-12th class | 2.18 (1.79, 2.64) | <0.001 | 2.27 (1.77, 2.92) | <0.001 |
| ≤5th class | 3.51 (2.78, 4.43) | <0.001 | 3.27 (2.47, 4.32) | <0.001 |
| Illiterate | 1.37 (1.05, 1.78) | 0.022 | 1.48 (1.06, 2.05) | 0.020 |
| Husband's occupation | | | | |
| Professional, managerial and technical | 1.00 | | 1.00 | |
| Skilled manual and non-manual | 0.92 (0.69, 1.24) | 0.591 | 0.64 (0.46, 0.89) | 0.008 |
| Partly skilled and unskilled | 0.86 (0.63, 1.17) | 0.329 | 0.81 (0.57, 1.14) | 0.230 |
| Unemployed | 2.63 (1.76, 3.92) | <0.001 | 0.49 (0.31, 0.79) | 0.003 |
| **Induction/augmentation with non-/contra-indication** | | | | |
| Religion | | | | |
| Hindu | 1.00 | | 1.00 | |
| Muslim | 1.14 (0.93, 1.40) | 0.217 | 1.58 (1.28, 1.96) | <0.001 |
| Christian | 1.35 (0.89, 2.03) | 0.157 | 4.61 (3.12, 6.79) | <0.001 |
| Sikh/others | 0.26 (0.15, 0.44) | <0.001 | 0.45 (0.22, 0.94) | 0.033 |
| Residence | | | | |
| Rural | 1.00 | | 1.00 | |
| Suburb/urban | 1.97 (1.63, 2.37) | <0.001 | 1.43 (1.12, 1.83) | 0.004 |
| Living below poverty line (BPL) | | | | |
| not BPL | 1.00 | | 1.00 | |
| BPL certificate/self-certified | 1.71 (1.44, 2.02) | <0.001 | 2.51 (2.02, 3.13) | <0.001 |
| unknown | 0.43 (0.32, 0.57) | <0.001 | 0.47 (0.32, 0.67) | <0.001 |
| Woman's education | | | | |
| ≥12th class | 1.00 | | 1.00 | |
| 6-12th class | 1.21 (0.99, 1.47) | 0.061 | 1.31 (1.02, 1.68) | 0.034 |
| ≤5th class | 1.03 (0.79, 1.36) | 0.808 | 0.95 (0.69, 1.30) | 0.740 |
| Illiterate | 0.56 (0.40, 0.76) | <0.001 | 0.52 (0.35, 0.76) | <0.001 |
| Husband's occupation | | | | |
| Professional, managerial and technical | 1.00 | | 1.00 | |
| Skilled manual and non-manual | 0.70 (0.53, 0.94) | 0.018 | 0.45 (0.33, 0.62) | <0.001 |
| Partly skilled and unskilled | 0.66 (0.48, 0.91) | 0.011 | 0.53 (0.38, 0.74) | <0.001 |
| Unemployed | 0.49 (0.27, 0.89) | 0.018 | 0.09 (0.04, 0.19) | <0.001 |

‡Socio-economic factors mutually adjusted and additionally adjusted for age at labour, parity, body mass index in early pregnancy, gestational weight gain, previous pregnancy problems, pre-existing medical problems, number of antenatal check-ups, duration of iron-folic acid supplementation, healthcare professional managing childbirth and adverse lifestyles.
